# Racial/ethnic disparities in cardiovascular disease mortality attributable to long-term PM_2.5_ exposure in the United States from 2001 to 2016

**DOI:** 10.1101/2022.09.07.22279640

**Authors:** Yiqun Ma, Emma Zang, Ijeoma Opara, Yuan Lu, Harlan M. Krumholz, Kai Chen

## Abstract

**Background:** The average concentration of fine particulate matter (PM_2.5_) has decreased in the U.S. in recent years. However, the health benefits of this improvement among different racial/ethnic groups are not known. This study aimed to estimate the associations between long-term exposure to ambient PM_2.5_ and cause-specific cardiovascular disease (CVD) mortality rate and assess the PM_2.5_-attributable CVD deaths in non-Hispanic White, non-Hispanic Black, and Hispanic people across all counties in the contiguous U.S. from 2001 to 2016.

**Methods:** Using nationwide CVD mortality data for all ages obtained from National Center for Health Statistics, this study applied interactive fixed effects models to estimate the associations between 12-month moving average of PM_2.5_ concentrations and monthly age-adjusted CVD mortality rates by race/ethnicity, controlling for both measured and unmeasured spatiotemporal confounders. Mortality from major types of CVD (ischemic heart disease [IHD], myocardial infarction [MI], stroke, hypertensive disease, and hypertensive heart disease) was also studied. We then calculated the burden of PM_2.5_-attributable CVD deaths in different race/ethnicity groups and examined the magnitude of racial/ethnic disparity and its changes over time.

**Results:** A total of 13,289,147 CVD deaths were included in the study. Each 1-µg/m^3^ increase in 12-month moving average of PM_2.5_ concentration was associated with increases of 7.16 (95% confidence interval [CI]: 3.81, 10.51) CVD deaths per 1,000,000 Black people per month, significantly higher than the estimates for non-Hispanic White people (*P* value: 0.002). The higher vulnerability in non-Hispanic Black people was also observed for mortality from IHD, MI, and stroke. Long-term PM_2.5_ exposure contributed to approximately 75.47 (95% CI: 40.14, 110.80) CVD deaths per 1,000,000 non-Hispanic Black people annually, over 3 times higher than the estimated rate in non-Hispanic White people (16.89, 95% CI:13.17, 20.62). From 2001 to 2016, the difference in attributable CVD mortality rate between Black and White people reduced by 44.04% (from 75.80 to 42.42 per 1,000,000 people), but the burden in Black people was still over 3 times higher compared to White people.

**Conclusions:** Non-Hispanic Black people have the highest PM_2.5_-attributable CVD mortality burden. Although the racial/ethnic disparity in this burden was narrowed over time, the gap between racial/ethnic minorities and non-Hispanic White people remains substantial.

## Introduction

Ambient fine particulate matter (PM_2.5_) air pollution is a leading risk factor for mortality in the United States (U.S.), especially for mortality of cardiovascular diseases (CVD).^1^ The detrimental effect of PM_2.5_ on cardiovascular mortality persists even at concentrations below the current air quality standard.^2-5^ Over recent decades, PM_2.5_ levels have fallen considerably in the contiguous U.S., as a result of the national Clean Air Act and other regional efforts.^6, 7^ This improvement in air quality has yielded substantial benefits to human health.^8^ However, it is unclear whether the health benefits of this decreasing trend of PM_2.5_, especially to cardiovascular health, distributed equitably across different racial/ethnical groups.

Most previous studies examined this question focusing only on the PM_2.5_ exposure. Increasing evidence showed an unequal distribution of PM_2.5_ exposure across communities, with racial/ethnic minorities being exposed to disproportionately high levels of PM_2.5_.^9-14^ Despite the improvement in air quality during the past years, the disparity in PM_2.5_ exposure remains.^9, 11, 13, 14^ In the U.S., physical environment and neighborhoods tend to exhibit the symptoms of, and are shaped by, environmental racism.^15^ For example, racial/ethnic minorities in the U.S., such as Black and Hispanic people, tend to live in neighborhoods where they have higher exposure to air pollution,^14, 16^ close proximity to waste facilities,^17^ and low community green space (e.g. parks).^18, 19^ Such factors can all significantly increase the likelihood of PM_2.5_ exposure.

However, the racial/ethnic disparities in PM_2.5_-attributable mortality burden cannot be fully explained by the disparities in PM_2.5_ exposure. Different PM_2.5_-mortality relationships across racial/ethnic groups also need to be considered. Due to structural racism and its downstream social determinants, including lack of access to equitable healthcare resources and chronic racial stressors such as limited employment opportunities, safety concerns, exposure to racial discrimination, or over-policing,^20^ Black people may suffer from greater CVD mortality consequences of air pollution than White people, even when they live in the same physical environment.^21^ Therefore, it is important to use race/ethnicity-specific exposure-response function in health burden assessment.^22^

Utilizing the nationwide PM_2.5_ and cardiovascular mortality data, this study aimed to first estimate the associations of long-term exposure to PM_2.5_ with county-level cardiovascular mortality rates by race/ethnicity, and then assess the average annual PM_2.5_-attributable CVD mortality burden across racial/ethnic groups and how the racial/ethnic disparity in this burden changed over time between 2001 and 2016. Interactive fixed effects (IFE) models, a novel statistical method for causal inference, were applied to estimate the PM_2.5_-mortality exposure-response function. In addition to the total CVD mortality, we also analyzed mortality from major categories of CVD.

## Methods

### Study area and population

In total, this study covered 3,103 counties or county equivalents in the contiguous U.S., with consistent boundaries over the study period. The cartographic boundary for counties in the contiguous U.S. was downloaded from the U.S. Census Bureau’s TIGER/Line geodatabase.^23^ We merged counties with boundary changes from 2001 to 2016 with neighboring counties (**Supplemental Methods 1**). County-level population data was collected from the Surveillance, Epidemiology, and End Results Program, National Cancer Institute.^24^ We extracted the total population and population estimates by sex, age, race, and Hispanic origin for each county, 2001-2016.

### Mortality rate

Mortality data from 2001 to 2016 was provided by the National Center for Health Statistics, which included the year and month of death, the cause of death (International Statistical Classification of Diseases and Related Health Problems, 10^th^ Revision [ICD-10] codes), and the sex, age, race, and ethnicity of each deceased person. Information about the race and Hispanic origin of a decedent was based on death certificates. The categorization of race and ethnicity followed the national standards “Revisions to the Standards for the Classification of Federal Data on Race and Ethnicity”.^25^ This study focused on CVD mortality (ICD-10 code: I00-I99) and its major categories, including ischemic heart disease (IHD, I20-I25), stroke (I60-I69), and hypertensive diseases (HD, I10-I15). Deaths from more specific causes, myocardial infarction (MI, I21-I23) in IHD and hypertensive heart disease (HHD, I11) in HD, were also analyzed in this study.

We calculated the monthly county-level cause-specific mortality rates for male, female, non-Hispanic White, non-Hispanic Black, and Hispanic population. All mortality rates were age-adjusted by direct standardization using the 2000 U.S. Census population as the standard population. Mortality data in 2000 was also obtained to calculate the 12-month moving average of mortality rates in a sensitivity analysis.

Using anonymized monthly county-level mortality records, this study was determined as a Not Human Subject research and approved by the Yale Institutional Review Boards (protocol ID: 2000026808).

### Air pollution and temperature

Daily PM_2.5_ concentration data from 2000 to 2016 for the contiguous U.S. at a resolution of 1 km was obtained from the NASA Socioeconomic Data and Applications Center.^26^ In brief, the PM_2.5_ concentration was estimated by an ensembled model based on neural network, random forest, and gradient boosting, using satellite data, meteorological data, land-use variables, elevation, chemical transport model predictions, and other variables as predictors.^6^ The 10-fold cross-validated coefficients of determination (R^2^) of this model was 0.86 for daily predictions, indicating a strong predictive performance.^6^

The daily PM_2.5_ concentrations were aggregated to monthly county-level to match to the mortality data. For months from January 2001 to December 2016, we calculated the 12-month moving average of PM_2.5_ concentration for each county to represent the average exposure to PM_2.5_ in the previous one year.^27, 28^ Nitrogen dioxide (NO_2_) and ozone (O_3_) data was downloaded from the same source and processed in the same way as PM_2.5_.^29-32^

Monthly mean air temperature data at 4 km × 4 km was obtained from the PRISM Climate Group.^33^ Similar to the air pollution data, we averaged the air temperature data for each county.

### Statistical analysis

We applied IFE models to estimate the associations between long-term exposure to PM_2.5_ and cardiovascular mortality rates. The IFE model is an innovative method in economics for causal inference, which is more flexible than the commonly-used traditional two-way fixed effects (TWFE) model.^34^ TWFE models can potentially control for both spatial confounders that only vary across counties (time-invariant confounders) and temporal confounders that only vary by time (county-invariant confounders), either measured or unmeasured.^35^ However, this approach assumes no unmeasured confounders that display different temporal variations across counties (time-varying individual county effects), which is often violated.^36^ Therefore, we used an extension of the TWFE model, the novel IFE model, which can potentially account for unmeasured time-varying county-specific confounders.^34, 36, 37^ The IFE approach requires weaker assumptions than TWFE models: it only requires a correct functional form (a relaxed form of strict exogeneity) and the existence of a low-dimensional decomposition.^36^

In the main IFE model, the outcome variable was the monthly county-level mortality rate and the predictor of interest was the 12-month moving average of PM_2.5_ concentration for each county, each month. Time-invariant and county-invariant confounders were controlled for by indicator variables for each county and month and air temperature was controlled by a flexible natural cubic spline with five degrees of freedom (df). Unmeasured time-varying individual county effect was explicitly modelled by decomposition. Statistical details of the IFE model can be found in **Supplemental Methods 2**.

Based on the estimated coefficient of PM_2.5_ (*β*) for each cause, which can be interpreted as the increase in mortality rate associated with 1-µg/m^3^ increase in 12-month moving average of PM_2.5_ per month, we then calculated the PM_2.5_-attributable number of deaths (AN) in each county in each month by 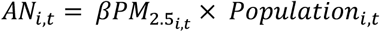, where 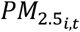 is the moving average of PM_2.5_ concentration of the current and previous 11 months for county *i*, month *t* and *Population*_*i,t*_ is the total population in county *i*, month *t*. The reference PM_2.5_ concentration was set as 0, since recent epidemiological evidence suggests that there is no safe level of PM_2.5_.^4,5^

We further estimated the association between long-term exposure to PM_2.5_ and cause-specific cardiovascular mortality rates and the attributable mortality burden by race/ethnicity (non-Hispanic White, non-Hispanic Black, and Hispanic people). We also conducted subgroup analysis by sex (male and female) to test sex differences and stratified analysis by urban or rural counties^38^ to investigate its potential modification effect. We tested the statistical difference in effect estimates between different groups by calculating the z score as 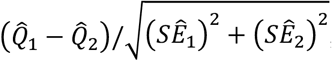, where 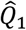 and 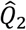 are the estimates, and *SÊ*_1_ and *SÊ*_2_ are their respective standard errors.^39^

Several sensitivity analyses were performed to test the robustness of our results: (a) counties with population size smaller than 5,000 or larger than 200,000 were excluded (approximately the 10^th^ and 90^th^ percentiles of county-level population size); (b) 12-month moving average of CVD mortality rates were used as the outcome; (c) traditional TWFE model was applied; (d) we additionally adjusted for NO_2_ or O_3_ in the model; (e) an alternative four or six df was used in the natural cubic spline of air temperature.

## Results

### Description of PM_2.5_ exposure and CVD mortality

A total of 13,289,147 CVD deaths was included in the study. From 2001 to 2016, the mean age-adjusted CVD mortality rate was 232.67 per 1,000,000 people per month in the 3,103 counties in the contiguous U.S. (standard deviation [SD]: 140.97), of which over 70% were from IHD, stroke, and HD. The mean age-adjusted CVD mortality rate was higher in males (278.43, SD: 230.46) than in females (195.85, SD: 166.79), and was the highest in non-Hispanic Black people (294.75, SD: 425.72) among the three studied racial/ethnical groups (**Table 1**). The mean 12-month moving average of PM_2.5_ concentration was 9.12 µg/m^3^ (SD: 3.13) (**Table 1**). The average PM_2.5_ concentrations were in general higher in the eastern U.S. (**Figure 1A**) and have greatly decreased in most counties in our study period (**Figure 1B**). Among people of different racial/ethnic groups who died from cardiovascular diseases from 2001 to 2016, although the long-term PM_2.5_ exposure reduced in all three groups (**Figure S1**), the median concentration of long-term PM_2.5_ exposure was the highest in the non-Hispanic Black group (**Figure 1C**).

**Table 1.** Monthly descriptive statistics for all 3,103 contiguous U.S. counties from 2001 to 2016^*^.

|  | Mean (SD<sup>†</sup>) | Min | Median (IQR<sup>‡</sup>) | Max |
| --- | --- | --- | --- | --- |
| *Environmental factors* |  |  |  |  |
| 12-month moving average of PM<sub>2.5</sub> (µg/m<sup>3</sup>) | 9.12 (3.13) | 1.54 | 9.23 (4.33) | 18.79 |
| Air temperature (°C) | 12.70 (9.90) | -20.58 | 13.57 (15.61) | 35.41 |
| *Age-adjusted mortality rate (per 1,000,000 individuals)* |  |  |  |  |
| Cardiovascular disease | 232.67 (140.97) | 0.00 | 217.52 (140.42) | 15,508.56 |
| Ischemic heart disease | 111.37 (97.71) | 0.00 | 97.88 (95.61) | 15,508.56 |
| Myocardial infarction | 47.70 (65.91) | 0.00 | 30.91 (66.48) | 4,169.58 |
| Stroke | 40.01 (53.16) | 0.00 | 30.48 (56.68) | 3,877.14 |
| Hypertensive disease | 14.32 (31.09) | 0.00 | 0.00 (18.90) | 2,584.76 |
| Hypertensive heart disease | 6.89 (21.80) | 0.00 | 0.00 (3.98) | 2,584.76 |
| *Age-adjusted CVD<sup>§</sup> mortality rate by race/ethnicity (per 1,000,000 individuals)* |  |  |  |  |
| Non-Hispanic White | 230.19 (158.33) | 0.00 | 213.54 (144.14) | 15,508.56 |
| Non-Hispanic Black | 294.75 (425.72) | 0.00 | 234.17 (407.62) | 44,840.80 |
| Hispanic | 134.11 (399.58) | 0.00 | 0.00 (141.02) | 22,011.77 |
| *Age-adjusted CVD mortality rate by sex (per 1,000,000 individuals)* |  |  |  |  |
| Male | 278.43 (230.46) | 0.00 | 250.13 (213.63) | 15,508.56 |
| Female | 195.85 (166.79) | 0.00 | 175.77 (151.65) | 15,508.56 |

\*Some neighboring counties were combined due to historical boundary changes
<sup>†</sup>SD: standard deviation
<sup>‡</sup>IQR: interquartile range
<sup>§</sup>CVD: cardiovascular disease

**Figure 1.**
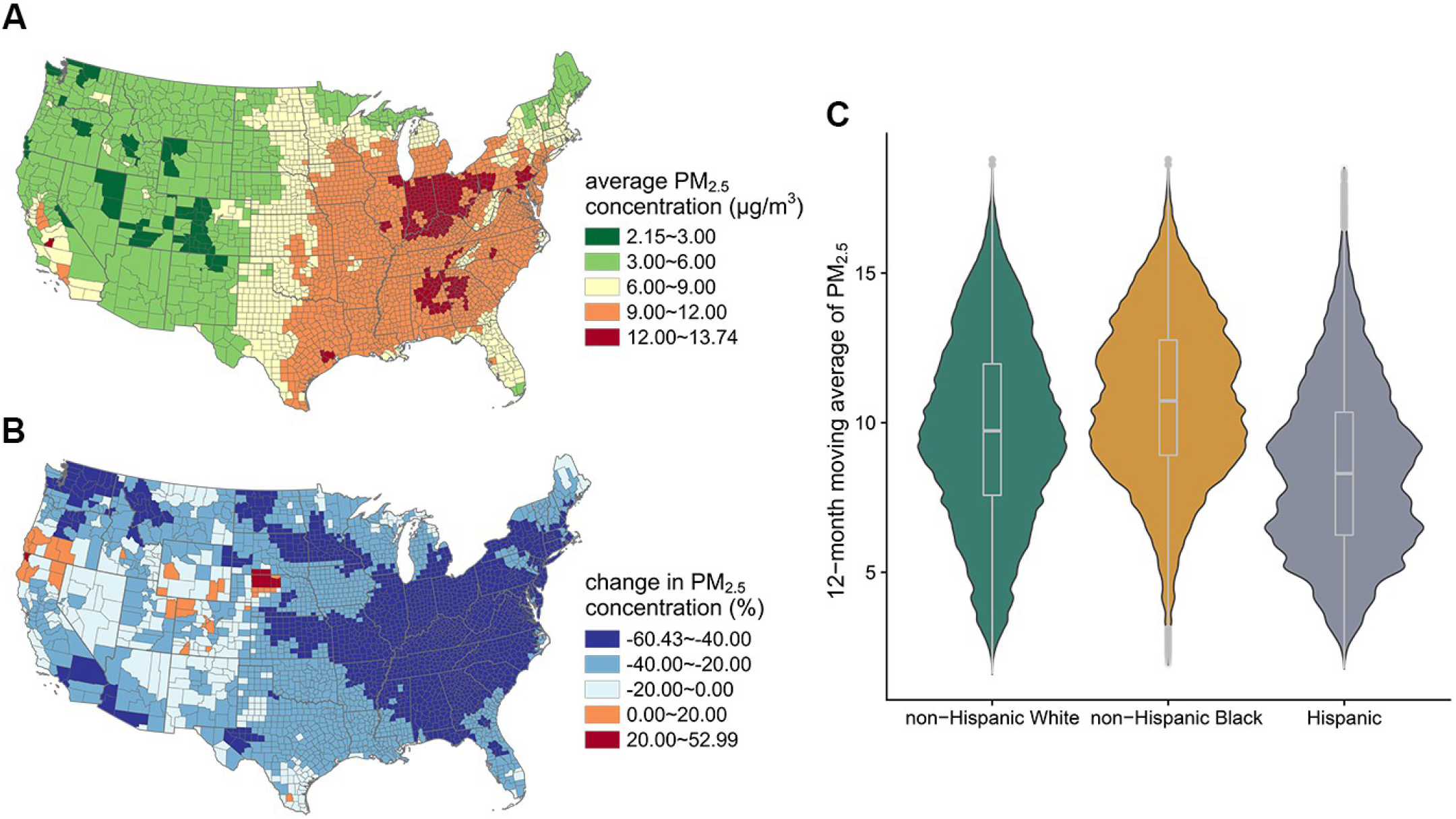
Distribution of PM_2.5_ concentration in all counties in the contiguous United States. **A**. Average monthly PM_2.5_ concentration from 2000 to 2016 in each county (µg/m^3^); **B**. Percent change in annual mean PM_2.5_ concentration from 2000 to 2016 (%); **C**. The distribution of long-term PM_2.5_ exposure (12-month moving average of PM_2.5_ concentration, µg/m^3^) among people of different racial/ethnic groups who died from cardiovascular diseases from 2001 to 2016

### Association between long-term PM_2.5_ exposure and cause-specific CVD mortality by race/ethnicity

We found that each 1-µg/m^3^ increase in the 12-month moving average of PM_2.5_ concentration was significantly associated with 2.01 additional CVD deaths per 1,000,000 people (95% confidence interval [CI]: 1.76, 2.35) per month. Compared to the non-Hispanic White group, the effect estimate was significantly higher in non-Hispanic Black group (*P* value: 0.002): each 1-µg/m^3^ increase in the 12-month moving average of PM_2.5_ concentration was significantly associated with 7.16 (95% CI: 3.81, 10.51) additional CVD deaths per 1,000,000 non-Hispanic Black people, while was only associated with 1.76 (95% CI: 1.37, 2.15) CVD deaths per 1,000,000 non-Hispanic White people (**Table 2**). No significant difference was observed between males and females (**Table S1**).

**Table 2.**
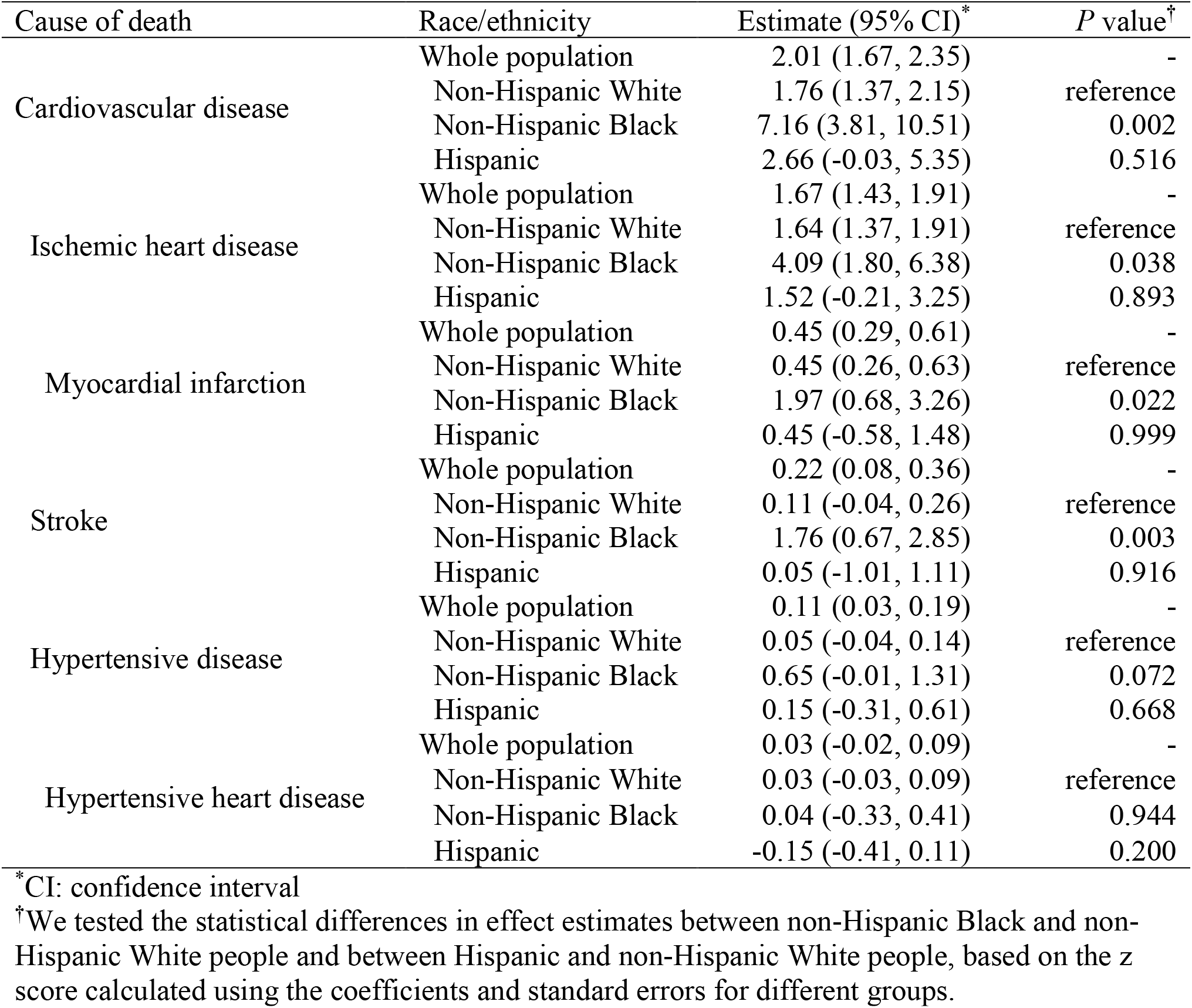
Deaths per 1,000,000 individuals associated with 1-µg/m^3^ increase in 12-month moving average of PM_2.5_ per month by race/ethnicity.

For major CVD categories, 1.67 (95% CI: 1.43, 1.91) IHD deaths, 0.22 (95% CI: 0.08, 0.36) stroke deaths, and 0.11 (95% CI: 0.11, 0.03, 0.19) HD deaths per 1,000,000 people were linked to a 1-µg/m^3^ increase in PM_2.5_. Higher long-term exposure to PM_2.5_ was also associated with higher mortality rates of MI and HHD. For mortality from IHD and its subtype MI, the effect estimates of the non-Hispanic Black group were the highest among all racial/ethnic groups (IHD: 4.09, 95% CI: 1.80, 6.38; MI: 1.97, 95% CI: 0.68, 3.26). In addition, the association between long-term PM_2.5_ exposure and stroke mortality rate was only significantly positive in the non-Hispanic Black group (**Table 2**).

### Average annual PM_2.5_-attributable CVD mortality burden by race/ethnicity

The average annual PM_2.5_-attributable CVD mortality burden was in general higher in eastern counties, especially in the Ohio Valley region and the Southeast region, which is consistent with the distribution of PM_2.5_ concentrations (**Figure 2A**). In total, approximately 69,675 (95% CI: 57,785, 81,565) CVD deaths were attributable to long-term PM_2.5_ exposure each year from 2001 to 2016, which contributed to about 7.84% of total CVD deaths in the contiguous U.S. (**Figure 2B**). Among specific causes, approximately 57,889 IHD deaths (including 15,634 MI deaths), 7,487 stroke deaths, and 3,882 HD deaths (including 1,192 HHD deaths) were attributable to PM_2.5_ each year (**Figure 2B; Table S2**).

**Figure 2.**
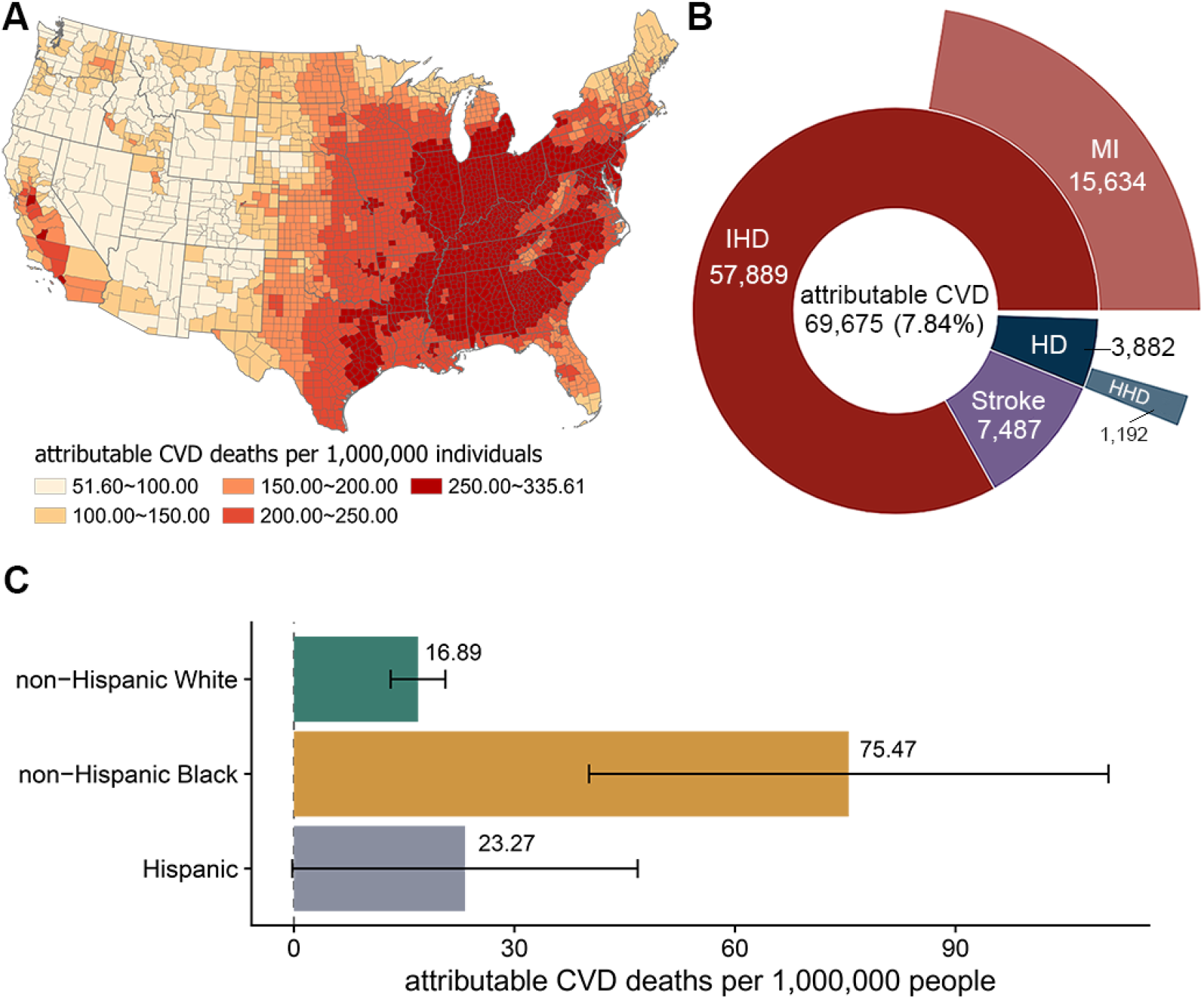
Average annual cardiovascular deaths attributable to PM_2.5_ (2001–2016) **A**. Average annual PM_2.5_-attributable cardiovascular deaths per 1,000,000 individuals from 2001 to 2016 in each county; **B**. Average annual PM_2.5_-attributable deaths from cardiovascular disease (CVD), ischemic heart disease (IHD), myocardial infarction (MI), stroke, hypertensive disease (HD), and hypertensive heart disease (HHD) from 2001 to 2016; **C**. Average annual PM_2.5_-attributable cardiovascular deaths per 1,000,000 individuals from 2001 to 2016 in each racial/ethnic group.

Among racial/ethnical groups, the average annual PM_2.5_-attributable CVD mortality burden was 40,315 (95% CI: 31.426, 49,205) deaths in non-Hispanic White people, 35,136 (95% CI: 18,689, 51,583) deaths in non-Hispanic Black people, and 13,335 (95% CI: -126, 26,795) deaths in Hispanic people. Although the absolute number of PM_2.5_-attributable deaths was the highest in the non-Hispanic White group, non-Hispanic Black people still shouldered the highest relative burden: in average, long-term PM_2.5_ exposure contributed to approximately 75.47 (95% CI: 40.14, 110.80) CVD deaths per 1,000,000 non-Hispanic Black people annually, over 3 times higher than the estimated rate in non-Hispanic White people (16.89, 95% CI:13.17, 20.62) (**Figure 2C**).

### Changes in racial/ethnical disparity in PM_2.5_-attributable CVD mortality burden over time

From 2001 to 2016, the CVD mortality burden attributable to long-term PM_2.5_ exposure has greatly decreased in most U.S. counties, especially the eastern U.S., with the greatest decline in West Virginia and Kentucky (**Figure 3A**). In total, PM_2.5_-attributable CVD deaths have decreased by 34.19%, from 82,047 (95% CI: 68,046, 96,048) in 2001 to 53,996 (95% CI: 44,782, 63,210) in 2016 (**Figure 3B; Table S3**).

**Figure 3.**
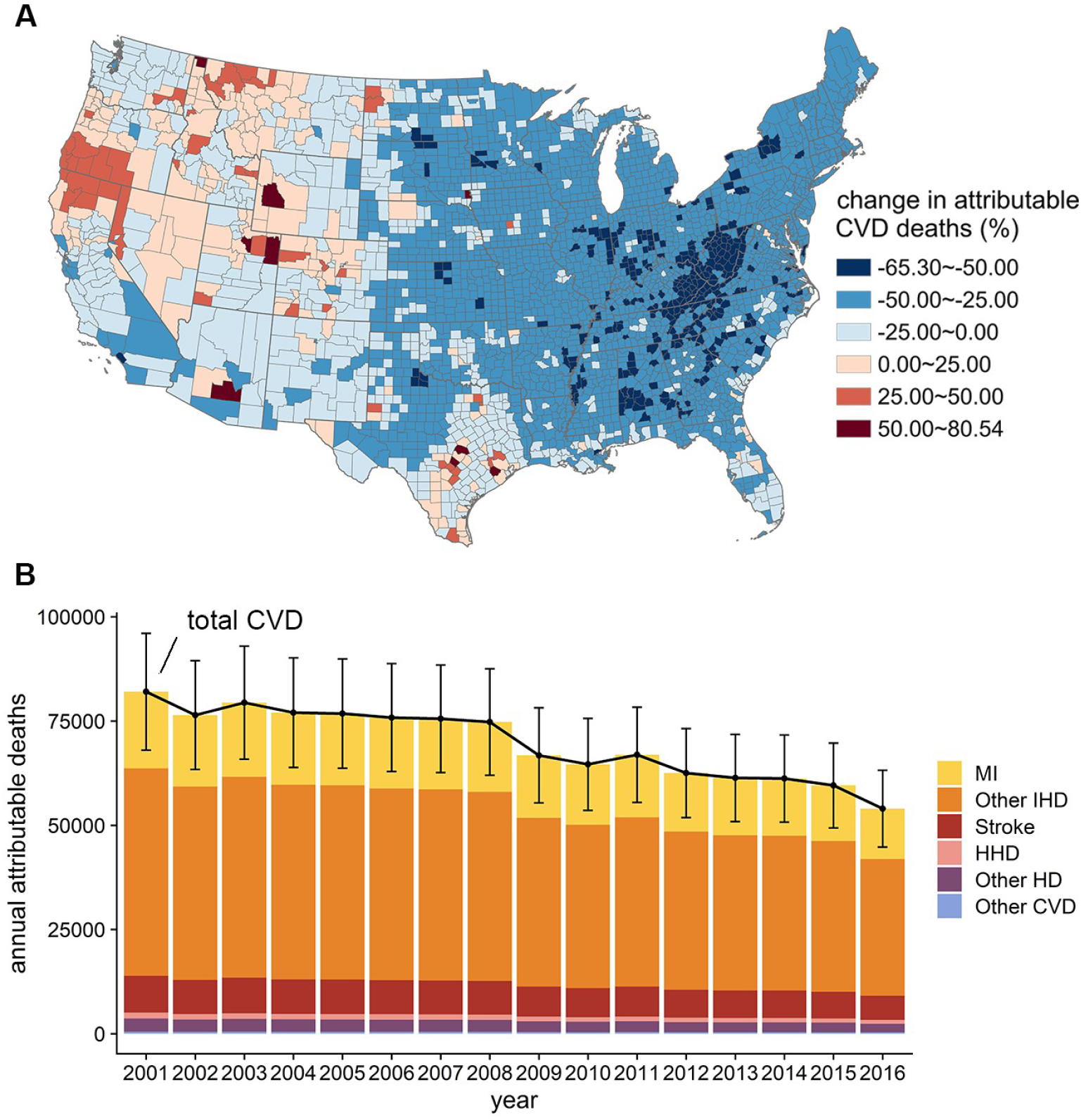
Changes in annual cardiovascular deaths attributable to PM_2.5_ by county and by specific cause from 2001 to 2016. **A**. Percent change in annual PM_2.5_-attributable cardiovascular deaths from 2001 to 2016 in each county; **B**. Annual PM_2.5_-attributable deaths from cardiovascular disease (CVD), including myocardial infarction (MI) and other ischemic heart disease (IHD), stroke, hypertensive heart disease (HHD) and other hypertensive disease (HD), and other CVD, from 2001 to 2016.

Despite of the overall decreasing trend, the magnitudes of decrease varied across different racial/ethnical groups (**Figure 4A**). The non-Hispanic Black group experienced the greatest reduction in PM_2.5_-attributable CVD mortality burden, from 96.90 deaths (95% CI: 51.54, 142.25) in 2001 to 54.65 (95% CI: 29.07, 80.23) in 2016, per 1,000,000 Black people. As a result, the racial/ethnic disparity became smaller: comparing to non-Hispanic White people, the PM_2.5_-attributable CVD mortality rate in non-Hispanic Black people was 75.80 higher in 2001 and 42.42 higher in 2016, which reduced by 44.04%; this difference also reduced by 29.62% for the Hispanic group, from 7.90 in 2001 to 5.56 in 2016 (**Figure 4B**).

**Figure 4.**
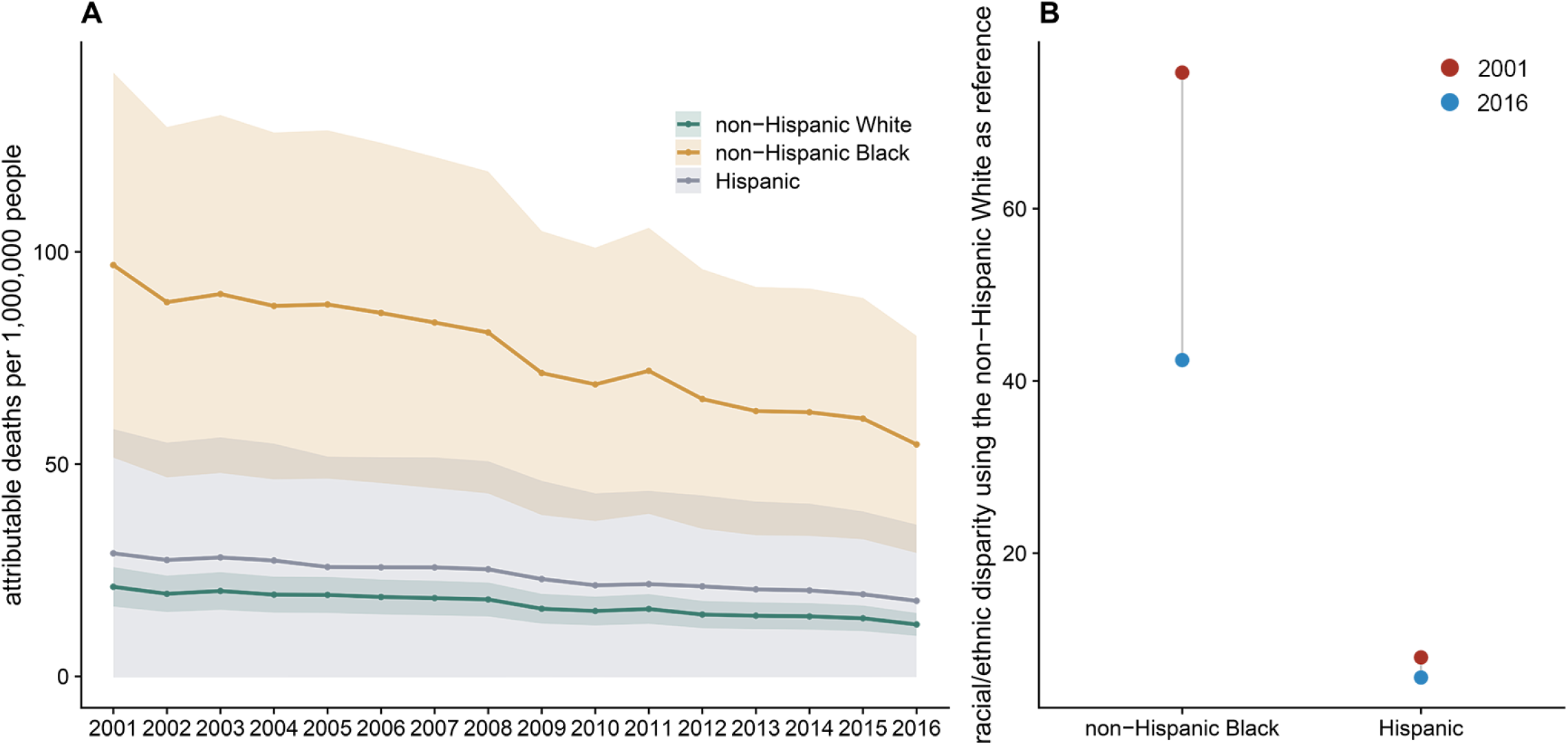
Racial/ethnic disparity in annual cardiovascular mortality rates attributable to PM_2.5_ from 2001 to 2016. **A**. Annual PM_2.5_-attributable cardiovascular disease (CVD) deaths per 1,000,000 people by race/ethnicity from 2001 to 2016; **B**. Racial/ethnic disparity in PM_2.5_-attributable CVD deaths per 1,000,000 people, using non-Hispanic White people as the reference group.

### Effect modification by urban vs. rural counties and sensitivity analyses

We observed a higher coefficient of PM_2.5_ in rural counties compared to urban counties. In rural counties, each 1-µg/m^3^ increase in long-term PM_2.5_ exposure was significantly associated with 2.05 CVD deaths per 1,000,000 people (95% CI: 1.54, 2.56) per month, while in urban counties, the effect estimate of PM_2.5_ became smaller (1.00, 95% CI: 0.62, 1.37). Consistent with the findings from our main analyses, in both rural and urban counties, the association between PM_2.5_ exposure and CVD mortality rate was stronger in non-Hispanic Black people compared to non-Hispanic White people (**Table S4**).

Sensitivity analyses showed that our results generally remained robust when excluding counties with population size smaller than 5,000 or greater than 200,000, using 12-month moving average of CVD mortality rate as the outcome, applying traditional TWFE, additionally adjusting for NO_2_ or O_3_ in the model, and using alternative numbers of df for air temperature (**Table S5**). The time-series plot of the monthly CVD mortality rates in counties with population size smaller than 5,000 indicated that the outcome measure was stable even in relatively small counties (**Figure S2**).

## Discussion

This study demonstrates significant racial/ethnic disparities in both vulnerability to and burden of PM_2.5_-related CVD mortality. First, the association between PM_2.5_ exposure and CVD mortality rate was significantly stronger for non-Hispanic Black people than non-Hispanic White people in the study. Second, non-Hispanic Black people also had the highest average annual PM_2.5_-attributable CVD mortality burden. Finally, the racial/ethnic disparities in this burden narrowed over time from 2001 to 2016, but the gap between racial/ethnic minorities and non-Hispanic White people persisted.

We found long-term exposure to ambient PM_2.5_ was associated with a greater increase in CVD mortality rate among non-Hispanic Black people compared to non-Hispanic White people. This finding from our study for all ages is consistent with the finding from a previous study focusing on people 65 years of age or older. A cohort study based on all Medicare beneficiaries reported that Black people had a higher estimated risk of all-cause mortality in association with PM_2.5_ exposure than the general population.^27^ This higher vulnerability of Black people can be explained by residential segregation and other downstream social determinants of structural racism in the U.S.^40, 41^ For example, Black people in the U.S. tend to have limited access to high-quality healthcare, education, credit, and other socioeconomic opportunities due to systemic racism. In addition, racism—intersecting and mutually reinforcing societal systems and institutions which foster and perpetuate racial discrimination and structure opportunities inequitably—in the U.S., contribute to higher levels of stress and likelihood of living in neighborhoods that are impacted by environmental racism, resulting in both higher CVD burden and vulnerability to air pollution.^22, 41^

In our study, the relative burden of PM_2.5_-attributable CVD mortality in non-Hispanic Black people was found to be over 3 times greater than the burden in non-Hispanic White people. The disparity in this annual burden between Black and White people (58.6 additional CVD deaths per 1,000,000 people) is of the same magnitude as the contribution of alcohol use to CVD mortality in the whole U.S. population in 2019 (55.3 CVD deaths per 1,000,000 people), according to the estimates from the GBD study.^1^ This finding reflects both disproportionally higher PM_2.5_ exposure and the aforementioned higher vulnerability of non-Hispanic Black people in the U.S. Multiple studies consistently demonstrated that people of color are typically exposed to higher levels of air pollution than non-Hispanic White people in the U.S,^9-12^ which is a pervasive and persistent consequence of redlining, inequitable siting of emission sources (e.g., highways and industrial facilities), and other policies and practices.^40, 41^

Benefited by the improvement of air quality in recent decades in the U.S., the overall PM_2.5_-attributable CVD mortality burden was greatly reduced. Among racial/ethnic groups, this burden decreased most among non-Hispanic Black people, thus narrowed the racial/ethnic disparity. However, this reduced disparity found in our study was mainly driven by the high vulnerability of Black people, not by a more equal distribution of air pollution. If we assume the exposure-response function of non-Hispanic Black people is the same as the one of non-Hispanic White people, the decreasing trends of PM_2.5_-attributable CVD mortality burden in these two groups would become almost parallel (**Figure S3**). Similarly, relying on a uniform relative risk estimate for the whole population, a recent preprint reported a widened gap between the most and least White communities regarding PM_2.5_-attributable premature deaths from 2010 to 2019.^42^ These different results indicate the importance of using race/ethnicity-specific exposure-response function when calculating the PM_2.5_-attributable mortality burden.^22^

Our results suggested an effect modification by urbanization level of residence, with stronger associations between PM_2.5_ and CVD mortality rates in rural counties compared to urban counties. A similar pattern was found for the association between PM_10_ and non-accidental mortality in Italy.^43^ One possible explanation of this finding is the different mixture compositions of PM_2.5_ in rural and urban counties,^44^ which may have different effects on cardiovascular mortality.^43, 45^

This study revealed the possible public health burden of structural racism through ambient air pollution in the U.S. Our findings could help inform environmental agencies to be more aware of the effects of environmental racism as a social justice issue. In addition, there is a need to better design and inform policies that efficiently reduce environmental inequity and protect vulnerable population in the pursuit of improving overall air quality. In addition, strategies aiming to reduce chronic racial stressors and ensure equal access to healthcare, education, and other resources are also urgently needed to reduce the racial/ethnic inequities in health burden attributable to air pollution such as PM_2.5_.

To the best of our knowledge, this study is one of the first studies that examined the changes in racial/ethnic disparity in PM_2.5_-attributable cardiovascular mortality burden over time using race/ethnicity-specific exposure-response function. In addition, we applied IFE model, an innovative causal inference method that controls for both measured and unmeasured time-varying county-level confounders, to estimate the association between long-term PM_2.5_ exposure and mortality from principal CVD types.

However, some limitations should be noted. First, this is a county-level ecological study, which is susceptible to ecological fallacy. We were also unable to take selective migration into account. Second, we did not specifically analyze for people of races/ethnicities other than non-Hispanic White people, non-Hispanic Black people and Hispanic people. Members of other races such as American Indians, Asians, and Pacific Islanders were not included in the race/ethnicity specific analyses due to lack of statistical power. Third, the potential measurement error in mortality data and misreport in race/ethnic categories may influence our results. Finally, we were unable to further explore the potential mechanism of the racial/ethnic disparity in PM_2.5_ attributable burden, which can be examined in depth in future studies.

## Conclusions

In conclusion, our study indicated that non-Hispanic Black people have the highest PM_2.5_-attributable CVD mortality burden among all racial/ethnical groups. Although the racial/ethnic disparity in this burden was narrowed in recent decades, the gap between people of color and non-Hispanic White people still exists. Policies and practices that aim to reduce both exposure and vulnerability to PM_2.5_ for racial/ethnical minorities, in addition to investigating the effects of structural racism on health, is needed.

## Data Availability

Air pollution data are available online at the NASA Socioeconomic Data and Applications Center (https://sedac.ciesin.columbia.edu/data/set/aqdh-pm2-5-concentrations-contiguous-us-1-km-2000-2016). Air temperature are available online at the PRISM Climate Group (https://prism.oregonstate.edu/). County-level population data are available online from the Surveillance, Epidemiology, and End Results Program, National Cancer Institute. Monthly mortality data from the National Center for Health Statistics are not publicly available.

## Acknowledgements

Dr. Zang received support from the National Institute on Aging (R21AG074238-01), the National Institute on Minority Health and Health Disparities (1R01MD017298-01), the Research Education Core of the Claude D. Pepper Older Americans Independence Center at Yale School of Medicine (P30AG021342), and the Institution for Social and Policy Studies at Yale University. Dr. Opara received support from the National Institutes on Health Director’s Early Independence Award (DP5OD029636; PI: Ijeoma Opara).

## Author Contributions

Y.M. conducted formal analyses and drafted the manuscript. K.C. conceived of and supervised the conduct of this study and edited the manuscript. E.Z. applied and managed the mortality dataset, supervised the statistical analysis, and revised the manuscript. I.O contributed to the writing and manuscript revision. Y.L. and H.K. contributed to the interpretation of results and manuscript revision. All authors reviewed and approved the final version of this manuscript.

