## Supplemental Method for "Racial/ethnic disparities in cardiovascular disease mortality attributable to long-term PM_2.5_ exposure in the United States from 2001 to 2016"

**Supplemental Methods 1.** The merging process for counties with boundary changes

**Supplemental Methods 2.** Interactive fixed effects model

**Figure S1.** Changes of long-term $PM_{2.5}$ exposure (12-month moving average of $PM_{2.5}$ concentration) among people of different racial/ethnic groups who died from cardiovascular diseases between 2001 and 2016

**Figure S2.** Monthly cardiovascular mortality rate in counties with population size smaller than 5,000 people

**Figure S3.** Annual $PM_{2.5}$-attributable cardiovascular deaths per 1,000,000 people by race/ethnicity from 2001 to 2016, assuming the same effect of $PM_{2.5}$ on mortality rate for all groups

**Table S1.** Cardiovascular deaths per 1,000,000 individuals associated with 1-µg/m$^3$ increase in 12-month moving average of $PM_{2.5}$ per month by sex

**Table S2.** Average annual cardiovascular deaths attributable to $PM_{2.5}$ by specific cause (2001–2016)

**Table S3.** Estimated mean cardiovascular deaths attributable to ambient $PM_{2.5}$ in each year from 2001 to 2016 (95% confidence interval)

**Table S4.** Cardiovascular deaths per 1,000,000 individuals per month associated with 1-µg/m3 increase in 12-month moving average $PM_{2.5}$ in urban and rural counties

**Table S5.** Results from sensitivity analyses

**Supplemental References**

### Supplemental Methods 1. The merging process for counties with boundary changes

The following U.S. contiguous counties with boundary changes from 2001 to 2016 were merged with neighboring counties:

- Broomfield County, Colorado (FIPS code: 08014) was created from parts of four Colorado counties: Adams (08001), Boulder (08013), Jefferson (08059), and Weld (08123). We merged these five counties into one in this study.
- Yellowstone National Park territory (30113) was merged into Gallatin (30031) and Park (30067) counties. We merged these three spatial units into one county equivalent.
- The status of the independent city of Bedford (51515) returned to a town within Bedford County (51019) in 2013. We merged this independent city to Bedford County in years before this change.

**Supplemental Methods 2. Interactive fixed effects model**

We used an interactive fixed (IFE) model to estimate the associations between long-term exposure to $PM_{2.5}$ and cardiovascular mortality rates. The IFE model can potentially account for unmeasured time-varying county-specific confounders by decomposing them into heterogeneous impacts of common trends.[1]

In our study, the model can be expressed as

$$Mortality\ rate_{i,t} = \mu + \alpha_i + \theta_t + \beta PM_{2.5_{i,t}} + ns(Temperature_{i,t}, df = 5) + v_{i,t} + \varepsilon_{i,t},$$

in which

$$v_{i,t} = \sum_{l=1}^{d} \lambda_{i,l} f_{l,t}.$$

$Mortality\ rate_{i,t}$ is the cause-specific mortality rate in county $i$, month $t$ and $PM_{2.5_{i,t}}$ is the moving average of $PM_{2.5}$ concentration of the current and previous 11 months for county $i$, month $t$. $\alpha_i$ refers to time-invariant county effects and $\theta_t$ refers to time-varying effects that are common in all counties. Air temperature was controlled by a flexible natural cubic spline with five degrees of freedom (df). $v_{i,t}$ is the unmeasured time-varying individual county effect, which is decomposed into $d$ common time-varying factors $f_{l,t}$, with corresponding unobserved county-level loading parameters $\lambda_{i,l}$.[2, 3]

We selected the number of common time-varying factors $f_{l,t}$ in the interactive fixed effects model following the criteria proposed by Bai and Ng (2002).[2] The number of factors $\hat{d}$ can be obtained by minimizing the following criterion:

$$PC(l) = \frac{1}{nT}\sum_{i=1}^{n}\sum_{t=1}^{T}(y_{i,t} - \hat{y}_{i,t}(l))^2 + l g_{n,T}$$

for all $l \in \{1,2,\dots\}$, where $n$ is the number of counties, $T$ is the number of months, and $\hat{y}_{i,t}(l)$ is the fitted value for a given factor dimension $l$. $g_{n,T}$ is a penalty term, which penalizes the undesired variance reduction caused by an increasing number of factors $\hat{d}$. This penalty term can be estimated by

$$g_{n,T} = \hat{\sigma}^2 \frac{(n+T)}{nT} \log\left(\frac{nT}{n+T}\right)$$

where $\hat{\sigma}^2$ is the sample variance estimator of the residuals $\varepsilon_{i,t}$. This variance estimator $\hat{\sigma}^2$ can be obtained by

$$\hat{\sigma}^2(d_{max}) = \frac{1}{nT}\sum_{i=1}^{n}\sum_{t=1}^{T}(y_{i,t} - \hat{y}_{i,t}(d_{max}))^2$$

where $d_{max}$ is an arbitrary maximal dimension of factors that is greater than $d$. In our study, we set $d_{max}$ as 14, the square root of the number of included months (192 months from 2001 to 2016). All statistical analyses were conducted with R software (version 4.1.3) using the package *phtt*.[3]

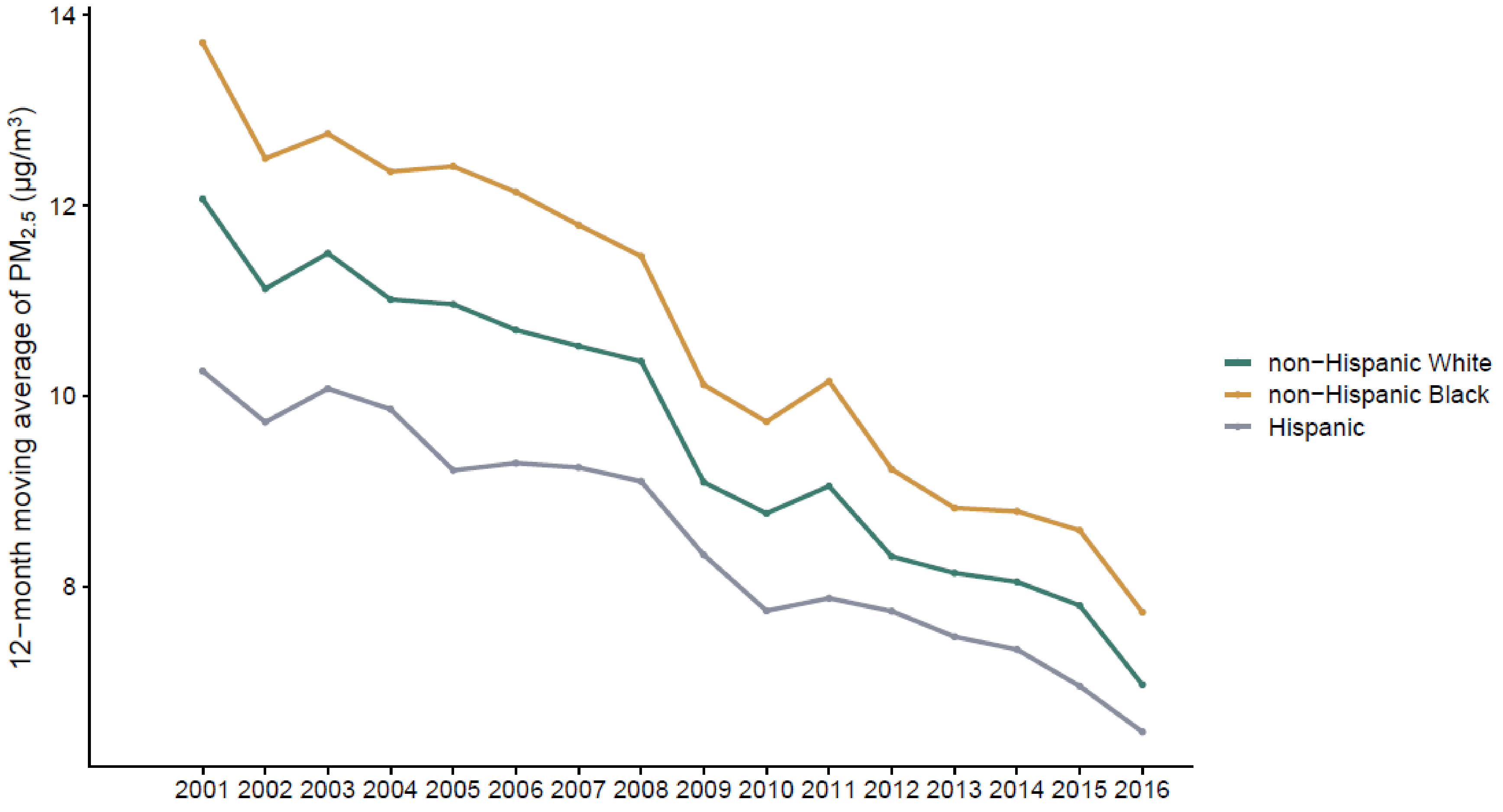


**Figure S1. Changes of long-term $PM_{2.5}$ exposure (12-month moving average of $PM_{2.5}$ concentration) among people of different racial/ethnic groups who died from cardiovascular diseases between 2001 and 2016**

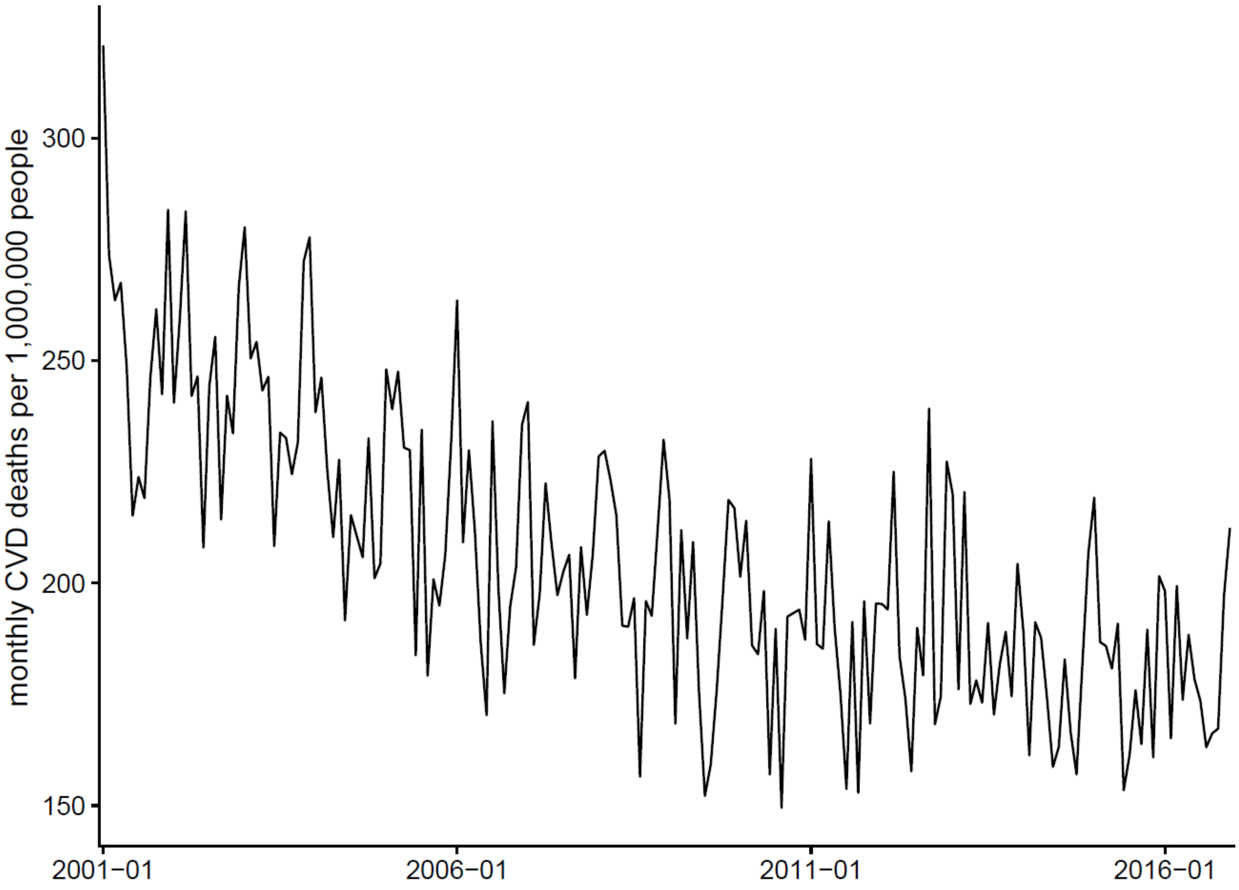


**Figure S2. Monthly cardiovascular mortality rate in counties with population size smaller than 5,000 people**

CVD: cardiovascular disease

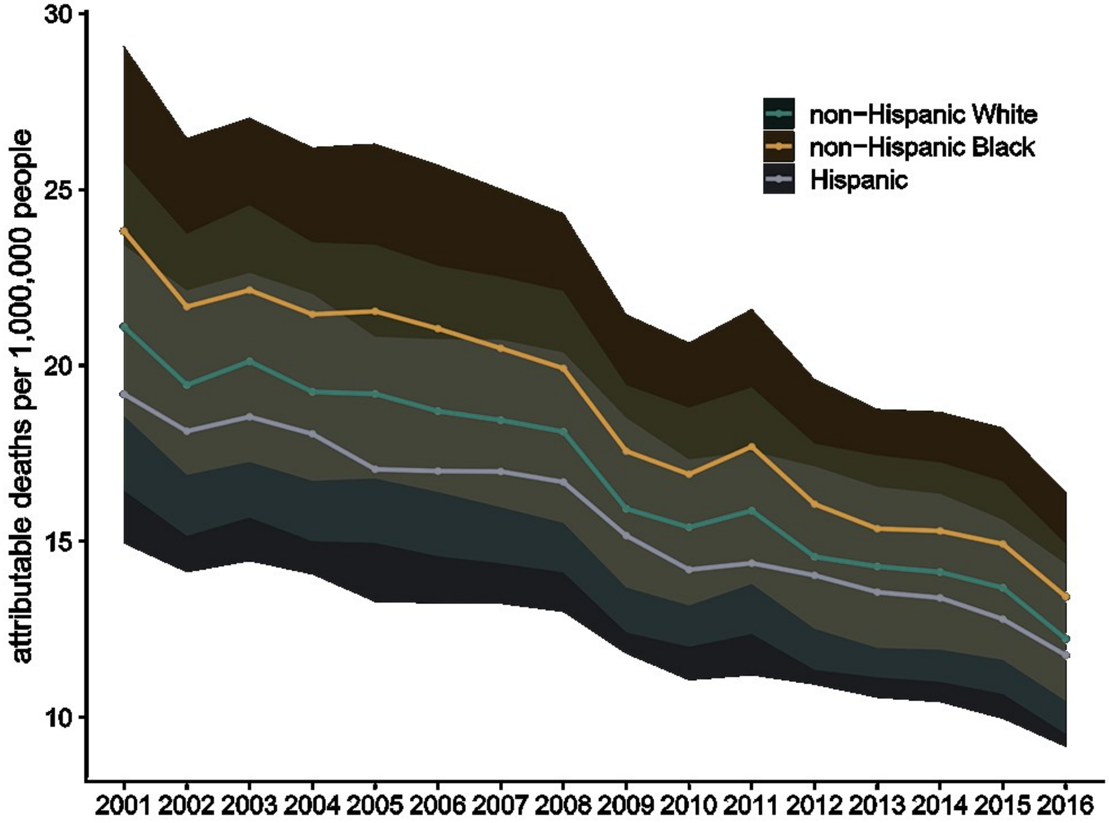


**Figure S3. Annual $PM_{2.5}$-attributable cardiovascular deaths per 1,000,000 people by race/ethnicity from 2001 to 2016, assuming the same effect of $PM_{2.5}$ on mortality rate for all groups**

This figure shows the time trend of $PM_{2.5}$-attributable cardiovascular disease mortality rates in different racial/ethnic groups using the exposure-response function of non-Hispanic White people for all groups. The comparison between this figure and Figure 4A indicates that the high vulnerability of non-Hispanic Black people is the main driver of the reduced racial/ethnic disparity observed in our study.

**Table S1. Cardiovascular deaths per 1,000,000 individuals associated with 1-μg/m$^3$ increase in 12-month moving average of $PM_{2.5}$ per month by sex**

| | Estimate (95% CI)* | *P* value† |
|---|---|---|
| Male | 2.33 (1.74, 2.92) | 0.357 |
| Female | 1.99 (1.57, 2.41) | |

*CI: confidence interval

†We tested the statistical difference in effect estimates between males and females based on the z score calculated using the coefficients and standard errors for different sexes.

**Table S2. Average annual cardiovascular deaths attributable to $PM_{2.5}$ by specific cause (2001–2016)**

| Cause of deaths | Attributable deaths |
|---|---|
| Cardiovascular disease | 69,675 (57,785, 81,565) |
| Ischemic heart disease | 57,889 (49,668, 66,110) |
| Myocardial infarction | 15,634 (10,001, 21,266) |
| Stroke | 7,487 (2,664, 12,311) |
| Hypertensive disease | 3,882 (1,110, 6,654) |
| Hypertensive heart disease | 1,192 (-724, 3,108) |

**Table S3. Estimated mean cardiovascular deaths attributable to ambient $PM_{2.5}$ in each year from 2001 to 2016 (95% confidence interval)**

| Year | Cardiovascular disease | Ischemic heart disease | Myocardial infarction | Stroke | Hypertensive disease | Hypertensive heart disease |
|---|---|---|---|---|---|---|
| 2001 | 82,047 (68,046, 96,048) | 68,169 (58,488, 77,849) | 18,410 (11,777, 25,042) | 8,817 (3,137, 14,497) | 4,572 (1,308, 7,836) | 1,404 (-852, 3,660) |
| 2002 | 76,426 (63,384, 89,468) | 63,498 (54,481, 72,516) | 17,148 (10,970, 23,326) | 8,213 (2,922, 13,504) | 4,259 (1,218, 7,299) | 1,308 (-794, 3,410) |
| 2003 | 79,426 (65,872, 92,980) | 65,991 (56,619, 75,362) | 17,821 (11,401, 24,242) | 8,535 (3,036, 14,034) | 4,426 (1,266, 7,586) | 1,359 (-825, 3,543) |
| 2004 | 77,015 (63,872, 90,157) | 63,987 (54,900, 73,074) | 17,280 (11,055, 23,506) | 8,276 (2,944, 13,608) | 4,291 (1,227, 7,355) | 1,318 (-800, 3,436) |
| 2005 | 76,782 (63,680, 89,885) | 63,794 (54,735, 72,854) | 17,228 (11,021, 23,435) | 8,251 (2,935, 13,567) | 4,278 (1,224, 7,333) | 1,314 (-797, 3,425) |
| 2006 | 75,829 (62,889, 88,769) | 63,002 (54,055, 71,949) | 17,014 (10,884, 23,144) | 8,149 (2,899, 13,399) | 4,225 (1,208, 7,242) | 1,298 (-787, 3,383) |
| 2007 | 75,545 (62,654, 88,437) | 62,766 (53,853, 71,680) | 16,951 (10,844, 23,058) | 8,118 (2,888, 13,349) | 4,209 (1,204, 7,215) | 1,293 (-784, 3,370) |
| 2008 | 74,773 (62,013, 87,533) | 62,125 (53,302, 70,947) | 16,777 (10,733, 22,822) | 8,035 (2,858, 13,212) | 4,166 (1,192, 7,141) | 1,280 (-776, 3,336) |
| 2009 | 66,769 (55,375, 78,163) | 55,475 (47,597, 63,353) | 14,982 (9,584, 20,379) | 7,175 (2,553, 11,798) | 3,720 (1,064, 6,377) | 1,143 (-693, 2,979) |
| 2010 | 64,615 (53,588, 75,641) | 53,685 (46,061, 61,309) | 14,498 (9,275, 19,721) | 6,944 (2,470, 11,417) | 3,600 (1,030, 6,171) | 1,106 (-671, 2,883) |
| 2011 | 66,914 (55,495, 78,333) | 55,595 (47,700, 63,490) | 15,014 (9,605, 20,423) | 7,191 (2,558, 11,823) | 3,729 (1,066, 6,391) | 1,145 (-695, 2,985) |
| 2012 | 62,534 (51,862, 73,205) | 51,956 (44,577, 59,334) | 14,031 (8,976, 19,086) | 6,720 (2,391, 11,049) | 3,484 (997, 5,972) | 1,070 (-649, 2,790) |
| 2013 | 61,349 (50,880, 71,819) | 50,972 (43,733, 58,211) | 13,765 (8,806, 18,725) | 6,593 (2,345, 10,840) | 3,418 (978, 5,859) | 1,050 (-637, 2,737) |
| 2014 | 61,218 (50,772, 71,665) | 50,863 (43,640, 58,086) | 13,736 (8,787, 18,685) | 6,579 (2,340, 10,817) | 3,411 (976, 5,847) | 1,048 (-636, 2,731) |
| 2015 | 59,560 (49,396, 69,723) | 49,485 (42,457, 56,512) | 13,364 (8,549, 18,179) | 6,400 (2,277, 10,524) | 3,319 (949, 5,688) | 1,019 (-618, 2,657) |
| 2016 | 53,996 (44,782, 63,210) | 44,862 (38,491, 51,233) | 12,115 (7,751, 16,480) | 5,803 (2,064, 9,541) | 3,009 (860, 5,157) | 924 (-561, 2,409) |

**Table S4. Cardiovascular deaths per 1,000,000 individuals per month associated with 1-μg/$m^3$ increase in 12-month moving average $PM_{2.5}$ in urban and rural counties[*]**

| Type | Race/ethnicity | Estimate (95% CI) [†] | *P* value[‡] |
|---|---|---|---|
| Urban (1156 counties) | Whole population | 1.00 (0.62, 1.37) | - |
| | Non-Hispanic White | 1.14 (0.73, 1.55) | reference |
| | Non-Hispanic Black | 3.66 (0.25, 7.07) | 0.150 |
| | Hispanic | -0.08 (-2.92, 2.77) | 0.407 |
| Rural (1947 counties) | Whole population | 2.05 (1.54, 2.56) | - |
| | Non-Hispanic White | 1.72 (1.15, 2.29) | reference |
| | Non-Hispanic Black | 5.96 (1.18, 10.74) | 0.084 |
| | Hispanic | 4.71 (0.89, 8.53) | 0.129 |

[*]The urban-rural classification was based on the 2013 NCHS Urban-Rural Classification Scheme for Counties. We classified all metropolitan counties as "urban counties" and nonmetropolitan counties as "rural counties".

[†]CI: confidence interval

[‡] We tested the statistical differences in effect estimates between non-Hispanic Black and non-Hispanic White people and between Hispanic and non-Hispanic White people, based on the z score calculated using the coefficients and standard errors for different groups.

**Table S5. Results from sensitivity analyses**

| Model | Model coefficient* | Attributable deaths† |
|---|---|---|
| Main model | 2.01 (1.67, 2.35) | 69,675 (57,785, 81,565) |
| Exclude counties with population < 5,000 (311 counties) | 1.80 (1.51, 2.09) | 62,261 (52,295, 72,226) |
| Exclude counties with population > 200,000 (329 counties) | 2.10 (1.72, 2.48) | 24,051 (19,674, 28,429) |
| 12-month moving average of CVD mortality rate | 2.18 (1.98, 2.38) | 75,568 (68,502, 82,634) |
| Two-way fixed effects model | 2.13 (1.79, 2.47) | 73,835 (61,877, 85,792) |
| Adjust for $NO_2$ | 2.01 (1.67, 2.35) | 69,675 (57,785, 81,565) |
| Adjust for $O_3$ | 2.04 (1.68, 2.40) | 70,715 (58,281, 83,148) |
| Temperature df=4‡ | 2.00 (1.66, 2.34) | 69,328 (57,438, 81,218) |
| Temperature df=6 | 2.00 (1.66, 2.34) | 69,328 (57,438, 81,218) |

*Cardiovascular deaths per 1,000,000 individuals associated with 1-μg/m$^3$ increase in 12-month moving average $PM_{2.5}$ per month

†Average annual cardiovascular deaths attributable to $PM_{2.5}$ from 2001 to 2016

‡df: degree of freedom
